# Network-based Translation of GWAS Findings to Pathobiology and Drug Repurposing for Alzheimer’s Disease

**DOI:** 10.1101/2020.01.15.20017160

**Authors:** Jiansong Fang, Pengyue Zhang, Quan Wang, Yadi Zhou, Chien-Wei Chiang, Rui Chen, Bin Zhang, Bingshan Li, Stephen J. Lewis, Andrew A. Pieper, Lang Li, Jeffrey Cummings, Feixiong Cheng

**Author notes:** Corresponding to Feixiong Cheng, Ph.D., Assistant Professor, Lerner Research Institute, Cleveland Clinic. Co-first authors Emials (F.C.) and (L.L).

## Abstract

Genome-wide association studies (GWAS) have identified numerous susceptibility loci for Alzheimer’s disease (AD). However, utilizing GWAS to identify high-confidence AD risk genes (ARGs) that can guide development of new therapeutics for patients suffering from AD has heretofore not been successful. To address this critical problem in the field, we have developed a genotype-informed, network-based methodology that interrogates pathogenesis to identify new therapeutics. When applied to AD, this approach integrates GWAS findings, multi-omics data from brain samples of AD patients and preclinical AD models, drug-target networks, and the human protein-protein interactome, along with large-scale patient database validation and *in vitro* mechanistic observations in human microglia cells. Through this approach, we identified 103 ARGs validated by various levels of pathobiological evidence in AD. Via network-based prediction and population-based validation, we then showed that pioglitazone usage is significantly associated with decreased risk of AD (hazard ratio (HR) = 0.895, 95% confidence interval [CI] 0.841-0.951, P = 3.97 × 10^−4^) in a retrospective case-control validation. Pioglitazone is a peroxisome proliferator-activated receptor agonist used to treat type 2 diabetes, and propensity score matching cohort studies confirmed its association with reduced risk of AD in comparison to glipizide (HR =0.921, 95% CI 0.861-0.983, *P* = 0.0146), an insulin secretagogue that is also used to treat type 2 diabetes. *In vitro* experiments showed that pioglitazone downregulated glycogen synthase kinase 3 beta (GSK3β) and cyclin-dependent kinase (CDK5) in human microglia cells, supporting a possible mechanism-of-action for its beneficial effect in AD. In summary, we present an integrated, network-based methodology to rapidly translate GWAS findings and multi-omics data to genotype-informed therapeutic discovery in AD.

## Introduction

Alzheimer’s disease (AD) is a chronic neurodegenerative disorder associated with progressive cognitive decline, extracellular amyloid plaques, intracellular neurofibrillary tangles, and neuronal death (Masters *et al*., 2015, Long and Holtzman, 2019). AD and other dementias are an increasingly important global health burden, recently estimated to affect 43.8 million people worldwide (Nichols *et al*., 2019). Although genome-wide association studies (GWAS) have identified over 40 genome-wide significant susceptibility loci for AD (Lambert *et al*., 2013, Cuyvers and Sleegers, 2016, Jung *et al*., 2018, Jansen *et al*., 2019), translating these findings into identification of high-confidence AD risk genes (ARGs) and potential therapies has eluded the field. Indeed, since Dr. Alois Alzheimer first described the condition in 1906, scientists have not developed any effective disease modifying treatments (Alzheimer, 1907, Long and Holtzman, 2019).

The number of AD patients is expected to rise to 16 million by 2050 in the United States (U.S.) alone (Kodamullil *et al*., 2017, Alteri and Guizzaro, 2018), while the attrition rate for AD clinical trials (2002-2012) is estimated at 99.6% (Cummings *et al*., 2014). One possible explanation for why most candidate drugs fail in later-stage clinical trials is poor target selection. Broadly in disease, drug targets with genetic support have carried a high success rate amongst U.S. Food and Drug Administration (FDA)-approved therapies (Cook *et al*., 2014, Nelson *et al*., 2015). However, this has not been the case with AD, and the translational application of multi-omics data such as GWAS for target identification and therapeutic development in AD remains challenging.

We recently demonstrated the utility of network-based methodologies for accelerating target identification and therapeutic discovery by exploiting multi-omics profiles from individual patients in multiple complex diseases, including cardiovascular disease (Cheng and Desai, 2018), cancer (Cheng and Lu, 2019), and schizophrenia (Wang *et al*., 2019). We now posit that systematic identification of likely causal genes by incorporating GWAS findings and multi-omics profiles with human interactome network models will also reveal disease-specific targets for genotype-informed therapeutic discovery in AD. This approach entails unique integration of the genome, transcriptome, proteome, and the human protein-protein interactome. Here, we report on application of this process to AD (**Fig. 1A**).

**Figure 1.**
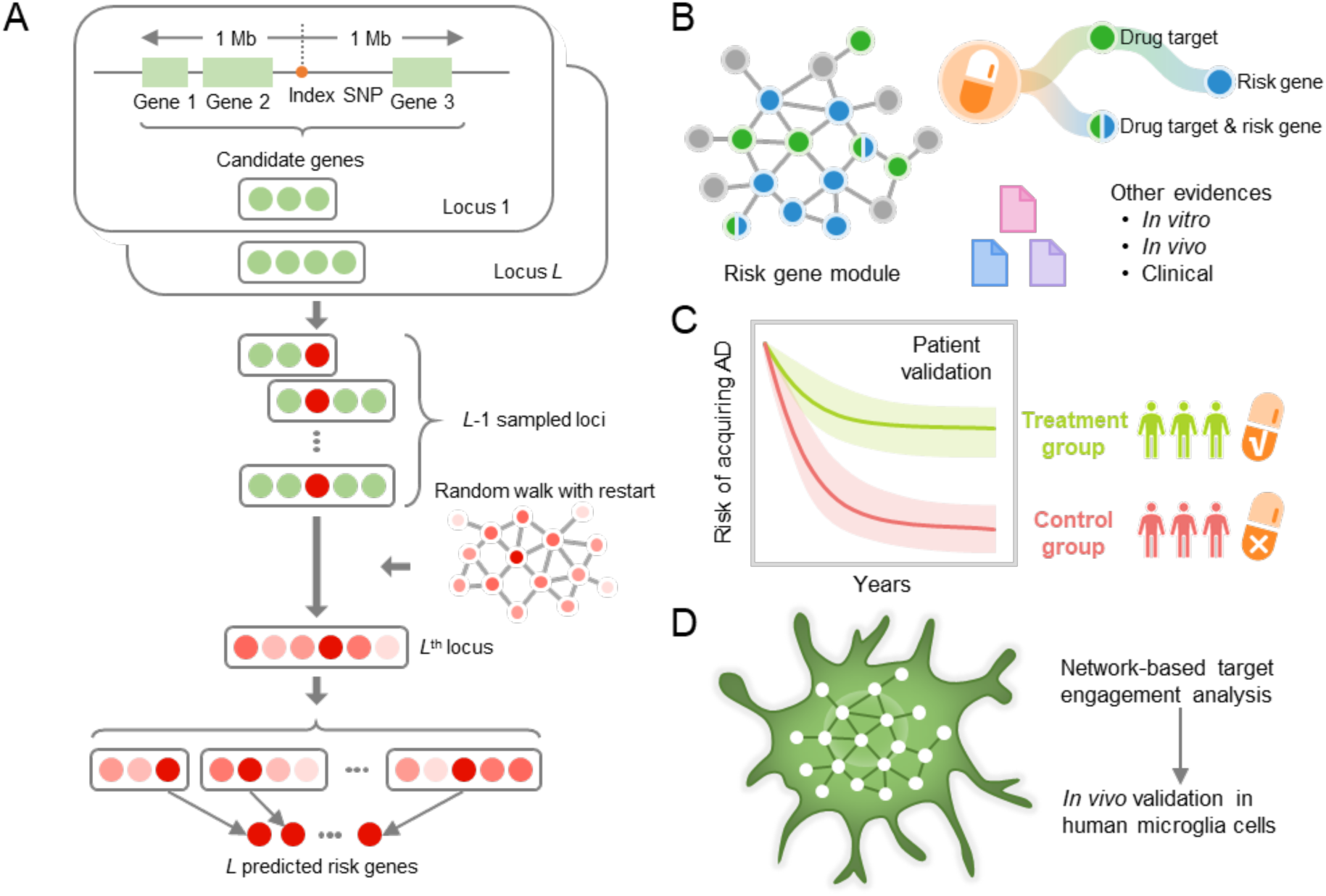
A diagram illustrating a genotype-informed, network methodology and population-based validation for Alzheimer’s therapeutic discovery. (**A**) A framework of network-based Bayesian algorithm (see Methods) for identifying Alzheimer’ disease (AD) risk genes. Specifically, this algorithm integrates multi-omics data and gene networks to infer risk genes from AD GWAS loci; (**B**) Network-based drug repurposing by incorporating ARGs and the human interactome network. (**C**) Population-based validation to test the drug user’s relationship with AD outcomes; Comparison analyses were conducted to evaluate the predicted drug-AD association based on individual level longitudinal patient data and the state-of-the-art pharmacoepidemiologic methods (see Methods). (**D**) Network-based mechanistic observation. Experimental validation of network-predicted drug’s proposed mechanism-of-action in human microglial cells.

Specifically, we integrated GWAS findings, multi-omics information generated from brain samples of individual AD patients and AD transgenic mouse models, publicly available drug-target networks, and the human protein-protein interactome, along with large-scale patient database validation and *in vitro* mechanistic observations in human microglia cells. By applying this network methodology to 106 AD loci, we identified a set of ARGs, most of which could be validated by multiple transcriptomic and proteomics profiles generated from AD transgenic mouse models. These ARGs represent enriched druggable targets for therapeutic discovery. Via a network-based prediction from findings of ARGs and population-based validation (**Fig. 1B and C**), we discovered that pioglitazone usage was significantly associated with decreased risk of AD in large-scale patient data. Subsequent *in vitro* mechanistic observations (**Fig. 1D**) revealed that pioglitazone significantly downregulates expression of glycogen synthase kinase 3 beta (GSK3β) and cyclin-dependent kinase 5 (CDK5) in human microglia cells, mechanistically supporting network-based and population-based findings.

## Materials and Methods

### Collection of GWAS SNPs from large-scale studies

In this study, we assembled multiple single nucleotide polymorphisms (SNPs) associated with AD from 15 large-scale GWAS studies in diverse population groups, conducted between 2007 and 2019 (**Supplementary Table 1**). Collectively, these studies include over 270,000 AD cases and 1100,000 controls. To maximize genetic signals based on the omnigenic hypothesis (Boyle *et al*., 2017), we adopted a loose threshold (P < 1 × 10^−5^) to collect AD risk SNPs, which yielded 106 unique GWAS SNPs.

### Construction of human protein-protein interactome

To build a comprehensive human protein-protein interactome, we assembled data from 15 common resources with multiple levels of experimental evidence. Specifically, we focused on high-quality protein-protein interactions (PPIs) with the following five types of experimental data: (1) binary PPIs tested by high-throughput yeast-two-hybrid (Y2H) systems; (2) kinase-substrate interactions by literature-derived low-throughput and high-throughput experiments; (3) literature-curated PPIs identified by affinity purification followed by mass spectrometry (AP-MS), Y2H and by literature-derived low-throughput experiments, and protein three-dimensional structures; (4) signaling network by literature-derived low-throughput experiments; (5) protein complex data (see **Supplementary Methods**). The genes were mapped to their Entrez ID based on the NCBI database, and duplicated pairs were removed. Collectively, the integrated human interactome included 351,444 PPIs connecting 17,706 unique proteins. More details are provided in our recent studies (Cheng and Desai, 2018, Cheng and Lu, 2019)

### Collection of functional genomics data

We collected the distal regulatory element (DRE)-promoter links inferred from two studies. The first study was the capture Hi-C study of cell line GM12878 (Mifsud *et al*., 2015). We obtained 1,618,000 DRE-promoter links predicted for GM12878 from http://www.ebi.ac.uk/arrayexpress/experiments/E-MTAB-2323/. The other dataset we used was from the FANTOM5 project (Andersson *et al*., 2014), in which cap analysis of gene expression (CAGE) technology was employed to infer enhancer-promoter links across multiple human tissues. We downloaded FANTOM5 data from http://enhancer.binf.ku.dk/presets/ and obtained 66,899 enhancer-promoter links.

### Collection of biological and functional data

#### Disease-associated genes from Open targets

Open targets refers to a comprehensive platform for therapeutic target identification and validation (Koscielny *et al*., 2017). We collected 527 AD disease-associated genes (**Supplementary Table 2**) from the Open Targets database (Assess in September, 2019).

#### Experimentally validated genes for Alzheimer’s disease

We further collected 144 high-quality experimentally validated AD genes (**Supplementary Table 2**), which combined genes involved in pathobiology of amyloidosis, tauopathy, or both, and genes characterizing other AD pathological hypothesis including neuroinflammation and vascular dysfunction (**Supplementary Method**).

#### Brain specific expression

We downloaded RNA-Seq data (RPKM value) of 32 tissues from GTEx V6 release (accessed on April 01, 2016, https://gtexportal.org/home/). We defined those genes with RPKM≥1 in over 80% of samples as tissue-expressed genes and the other genes as tissue-unexpressed. To quantify the expression significance of tissue-expressed gene *i* in tissue *t*, we calculated the average expression ⟨*E*(*i*)⟩ and the standard deviation *δ*_*E*_(*i*) of a gene’s expression across all considered tissues. The significance of gene expression in tissue *t* is defined as

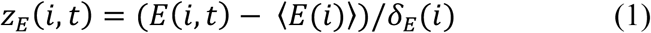

The details have been described in previous studies (Cheng and Desai, 2018, Cheng and Lu, 2019).

#### Gene Expression

We collected human microarray data in AD cases versus controls with brain samples from two independent datasets (GSE29378 and GSE84422) (Miller *et al*., 2013, Wang *et al*., 2016). We also collected mouse microarray data from AD transgenic mouse vs. controls, including brain microglia of 5XFAD mice from 2 independent datasets (GSE65067 and GSE74615) (Orre *et al*., 2014, Wang *et al*., 2015), and brain hippocampus of Tg4510 mice (GSE53480 and GSE57583) (Polito *et al*., 2014).

The original microarray datasets were obtained from Gene Expression Omnibus (https://www.ncbi.nlm.nih.gov/geo). Detailed information of these 6 GEO datasets is provided in **Supplementary Table 3**. All raw expression data were log2 transformed, and all samples were quantile normalized together. Probe IDs in each dataset were mapped to National Center for Biotechnology Information (NCBI) Entrez IDs, and probes mapping to multiple genome regions or without corresponding entrez IDs were deleted. The items were imported to R statistical processing environment using a LIMMA/Bioconductor package. All the mouse genes were further transferred into unique human-orthologous genes using the Mouse Genome Informatics (MGI) database (Eppig *et al*., 2017). Genes with threshold fold change (FC) > 1.2 were defined as exhibiting differential expression and prioritized as predicted AD risk genes.

### Enrichment analysis

Differentially expressed gene/protein (DEG/DEP) sets from multiple data sources were collected for enrichment analysis. This included a total of 6 bulk RNA-seq datasets and 10 proteomic datasets from 4 types of AD transgenic mouse models, including 5XFAD, Tg4510, ADLPAPT and hAPP (see **Supplementary Method**).

#### Bulk RNA-seq

We collected 2 RNA-seq datasets from brain or brain microglia of 5XFAD mice. In addition, we obtained 4 RNA-seq datasets from brain microglia of Tg4510 mice across different months [M] age (2M, 4M, 6 M, and 8M). Differential expression analysis was performed using DESeq (Anders, 2010), while threshold for significance of differential expression was set to FDR < 0.05 using Benjamini-Hochberg’s method. After mapping mouse genes to human-orthologous gene (Eppig *et al*., 2017), we obtained 6 differentially expressed gene sets.

#### Proteomics

In total, 10 proteomic datasets were assembled from 3 types of AD transgenic mouse models in two recent publications (Savas *et al*., 2017, Kim *et al*., 2018). The first study performed global quantitative proteomic analysis in hAPP and hAPP-PS1 mouse models at young (3 month [M]) and old ages (12 M) (Savas *et al*., 2017). We obtained four sets of DEPs (hAPP_3M, hAPP_12M, hAPP-PS1_3M and hAPP-PS1_12M) after merging the DEPs from different brain regions. The second study performed quantitative proteomics to uncover molecular and functional signatures in the hippocampus of three types of transgenic mice (Kim *et al*., 2018). Two of these mouse lines, including ADLPAPT (4M, 7M, 10M) that carry three human transgenes (APP, PSEN1 and tau) and hAPP-PS1 (4M, 7M, 10M) mouse, were used in this study. After mapping mouse genes to human-orthologous gene (Eppig *et al*., 2017), we obtained 10 sets of DEPs.

### AD risk gene prediction

We utilized our recently developed Bayesian model selection method to predict ARGs (**Fig 1A**), based on the assumption that true risk genes are more densely connected with each other in a biological network (Wang *et al*., 2019). Specifically, we collected at most 20 genes in the 2Mb region centered at a GWAS index SNP as the candidates for that particular locus. Assigning *L* as the number of GWAS loci, and we then denoted a vector of genes with length *L*, each being from one of the *L* GWAS loci, as (*X*_*1*_, …, *X*_*L*_), and termed it as candidate risk gene set (CRGS). Assigning *N* to represent the biological network, we then calculated *P(X*_*1*_,*…, X*_*L*_|*N)*, and selected a CRGS with maximum posterior probability. Computationally, it is not feasible to enumerate all possible gene combinations, and we therefore adopted a Gibbs sampling algorithm to transition the problem into a single-dimensional sampling procedure. For example, when sampling the risk gene from candidates at the *L*-th locus, we assumed that the risk genes at all other *L*-1 loci had been selected, and the sampling probability for a gene at the *L*-th locus was computed as conditional on the *L-1* risk genes, based on its closeness to other *L-1* risk genes in the network. For each candidate gene *X*_*L*_ at the *L*-th locus, we assigned *M*_*1*_ to represent the event that *X*_*L*_ is the risk gene at locus *L, M*_*0*_ represent the event that *X*_*L*_ is not the risk gene at locus *L*, and *X*_*-L*_ to represent all the selected risk genes in the other *L*-1 loci. The Bayesian model selection can be depicted as

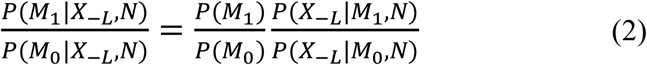

Where 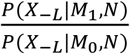 is a Bayesian Factor (BF) measuring the closeness between *X*_*-L*_ and *X*_*L*_ in network *N* and 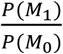 is prior odds. The prior odds reflect the prior knowledge whether *X*_*L*_ is a risk gene or not and we assumed *P(M*_*1*_) = *P(M*_*0*_) in this study. In regarding to 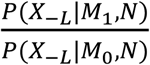, we adopted the random walk with restart (RWR) algorithm to calculate the BF. Starting from any node *n*_*i*_ in a predefined network *N*, the walker faces two options at each step: either moving to a direct neighbor with a probability 1 − *r* or jumping back to *n*_*i*_ with a probability *r*. The fixed parameter *r* is called the restart probability in RWR, and *r* was set as 0.3 in this study (Wang *et al*., 2019). Let *W* be the adjacency matrix that decides which neighbor to be moved to, and *q*_*t*_ be the reaching probability of all nodes at step *t*. The RWR algorithm is formalized as

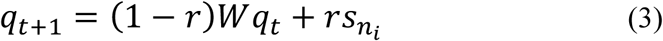

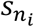 is a vector with the *i*-th element as 1 and 0 for others, which means th starting node is *n*_*i*_. Following the equation, *q*_*t*_ can be updated step by step until |*q*_*t*+1_ − *q*_*t*_|^2^ < *T*_*rwr*_, where *T*_*rwr*_ is a predefined threshold. We set *T*_*rwr*_ as 1e^-6^ (Wang *et al*., 2019). The adjacency matrix *W* represents the distance between any two nodes in the network and we adopted the same network and strategy in our previous work to calculate W. We calculated *P*(*X*_−*L*_|*M*_1_, *N*) based on *W*. We mapped *X*_*L*_ to the rows of *W* and *X*_−*L*_ to the columns of *W*, and obtained a vector with the same length as *X*_−*L*_. The sum of the vector was calculated as *P*(*X*_−*L*_|*M*_1_, *N*). In this study, we assumed *P*(*X*_−*L*_|*M*_0_, *N*) to be the same for all different candidate genes. Through the Bayesian model selection equation,

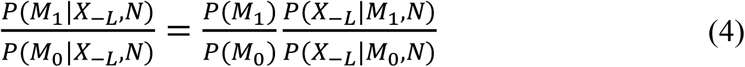

we obtained a value for each candidate genes at locus *L*. We used these values as sampling probabilities for Gibbs sampling to choose a risk gene for locus *L*. We then repeated the sampling across the remaining loci and iterated the sampling process until convergence. Specially, in each round of Gibbs sampling, we calculated the sampling frequency for each candidate gene. The frequency was compared with that of the previous round, and if the sum of squares of frequency differences across all selected genes was smaller than a predefined threshold (1 × 10^−4^ used in this study) then the sampling procedure was halted. Based on the sampling, we are able to assess the confidence of candidates being risk genes. In this study, we selected the gene with highest sampling frequency as risk gene for each locus as described previously (Wang *et al*., 2019).

### Construction of drug-target network

We integrated six commonly used resources to collect high-quality physical drug-target interactions for FDA-approved drugs. We obtained biophysical drug-target interaction using reported binding affinity data: inhibition constant/potency (*K*_*i*_), dissociation constant (*K*_*d*_), median effective concentration (*EC*_*50*_), or median inhibitory concentration (IC_50_) ≤ 10 µM. First, we extracted the bioactivity data from the DrugBank database (v4.3) (Wishart *et al*., 2018), the Therapeutic Target Database (TTD, v4.3.02) (Li *et al*., 2018), and the PharmGKB database (Barbarino *et al*., 2018). In addition, we acquired drug-target interactions related to FDA-approved drugs from three commonly used databases (Cheng and Lu, 2019). To improve data quality, we pooled only those items that satisfied the following four criteria: (i) binding affinities, including *K*_*i*_, *K*_*d*_, *IC*_*50*_, or *EC*_*50*_, ≤ 10 µM; (ii) the target protein has a unique UniProt accession number; (iii) proteins marked as “reviewed” in the UniProt database; and (iv) proteins are from Homo sapiens. Totally, we collected 15,367 drug– target interactions connecting 1,608 FDA-approved drugs and 2,251 unique human targets (**Supplementary Table 4**).

### Description of network proximity

Given the set of disease proteins (*A*), the set of drug targets (*B*), then the closest distance *d*_*AB*_ measured by the average shortest path length of all the nodes to the other module in the human protein-protein interactome can be defined as:

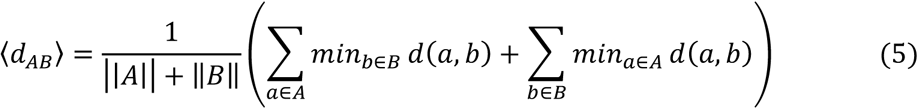

where *d*(*a, b*) denotes to the shortest path length between protein *a* and drug target *b*.

To calculate the significance of the network distance between a given drug and disease module, we constructed a reference distance distribution corresponding to the expected distance between two randomly selected groups of proteins of the same size and degree distribution as the original disease proteins and drug targets in the network. This procedure was run 1000 times. The mean 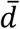 and standard deviation (*σ*_*d*_) of the reference distribution were used to caluculate a z-score (*z*_*d*_) by converting an observed (non-Euclidean) distance *d* to a normalized distance:

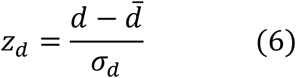

### Pharmacoepidemiologic validation

#### Patient Cohort Preparation

The pharmacoepidemiology study utilized the MarketScan Medicare Supplemental database from 2012 to 2017. The dataset included individual-level diagnosis codes, procedure codes, and pharmacy claims for ∼7 million patients per year. Pharmacy prescriptions of pioglitazone and glipizide were identified by using RxNorm and National Drug Code (NDC). For a subject exposed to the aforementioned drugs, a drug episode is defined as the time between drug initiation and drug discontinuation. Specifically, drug initiation is defined as the first day of drug supply (i.e. first prescription date). Drug discontinuation is defined as the last day of drug supply (i.e. last prescription date + days of supply) accompanied by no drug supply for the next 60 days. Gaps of less than 60-day of drug supply were allowed within a drug episode. The pioglitazone cohort included the first pioglitazone episode for each subject, as well as the glipizide cohort. Further, we excluded observations that started within 180-days of insurance enrollment. For the final cohorts, demographic variables including age, gender and geographical location were collected. Additionally, diagnoses of hypertension (HTN) and type 2 diabetics (T2D) (the ICD codes were given in **Supplementary Materials and Method**) before drug initiation were collected. These variables were specifically selected to address potential confounding biases. Lastly, a control cohort was selected from patients not exposed to pioglitazone. Specifically, non-exposures were matched to the exposures (ratio 1:4) by initiation time of pioglitazone, enrollment history, age, and gender. The geographical location, diagnoses of HTN and T2D were collected for the control cohort as well.

#### Outcome measurement

The outcome was time from drug initiation to diagnose of AD, which was defined by using the ICD codes (**Supplementary Materials and Method**). For pioglitazone and glipizide cohorts, observations without diagnose of AD were censored at the end of drug episodes. For the control cohort, the corresponding pioglitazone episode’s starting date was used as the starting time. Observations without diagnosis of AD were censored at the corresponding pioglitazone episode’s end date.

### Statistical analysis

Survival curves for time to AD were estimated using a Kaplan-Meier estimator. Additionally, propensity score stratified survival analysis was conducted to investigate the risk of AD between pioglitazone users and non-pioglitazone users, as well as pioglitazone users and glipizide users. For each comparison, the propensity score of taking pioglitazone was estimated by using a logistic regression model in which covariates included age, gender, geographical location, T2D diagnosis, and HTN diagnosis. Furthermore, propensity score stratified Cox-proportional hazards models were used to conduct statistical inference for the hazard ratios (HR) of developing AD between cohorts.

### Experimental validation

#### Reagents

Pioglitazone was acquired from Topscience. Lipopolysaccharides (LPS) (Cat# L2880) and 3-(4,5-dimethylthiazol-2-yl)-2,5-diphenyltetrazolium bromide (MTT) were obtained from Sigma-Aldrich. Antibodies against Phospho-GSK3B-Y216 (Cat# AP0261), GSK3B (Cat# A2081) and CDK5 (Cat# A5730), were purchased from ABclonal Technology. CDK5-Phospho-Tyr15 (Cat# YP0380) was obtained from Immunoway (Plano, Texas, USA). All other reagents were purchased from Sigma-Aldrich unless otherwise specified.

#### Cell viability

Human microglia HMC3 cells were purchased from American Type Culture Collection (ATCC, Manassas, VA). Cell viability was detected by MTT method as described previously (Huang *et al*., 2019). 5000 cells/well were plated in 96-well plates for 12 h, and then treated with pioglitazone for 48 h. After treatment, MTT solution was added to the cells to a final concentration of 1 mg/mL and the mixture was allowed to incubate at 37 °C for 4 h. The supernatant was removed, and precipitates were dissolved in DMSO. Absorbance was measured at 570 nm using a Synergy H1 microplate reader (BioTek Instruments, Winooski, VT, USA).

#### Western blot analysis

HMC3 cells were pre-treated with pioglitazone (3 µM or 10 µM) and DMSO (control vehicle), and followed with 1 µg/mL LPS for 30 min. Cells were harvested, washed with cold PBS, and then lysed with RIPA Lysis Buffer with 1% Protease Inhibitor (Cat# P8340, Sigma-Aldrich). Total protein concentrations were measured using a standard BCA protein assay kit (Bio-Rad, CA, USA), according to the manufacturer’s manual. Samples were electrophoresed by sodium dodecyl sulfate-polyacrylamide gel electrophoresis (SDS-PAGE), then blotted onto a polyvinylidene difluoride (PVDF; EMD Millipore, Darmstadt, Germany) membrane. After transferring, membranes were probed with specific primary antibodies (1:1000) at 4°C overnight. Specific protein bands were detected using a chemiluminescence reagent after hybridization with a horseradish peroxidase (HRP)-conjugated secondary antibody (1:3000).

## Results

### Pipeline of the network-based methodology

We utilized a Bayesian model selection method to predict ARGs (Wang *et al*., 2019), based on the assumption that likely causal risk genes are more densely connected with each other in a biological network (**Fig. 1A**). Specifically, we collected at most 20 genes in a 2 Mb region centered at a GWAS index SNP as the candidates for that particular locus. We then calculated a posterior probability for each candidate gene based on its closeness to other risk genes in the biological network. For each GWAS locus, the candidate gene with the highest posterior probability was predicted as an ARG (see Methods). By applying this framework to the 106 AD GWAS loci we collected (Methods), we predicted 103 ARGs after merging the overlapping genes across several different loci (**Supplementary Table 5**). Meanwhile, we also predicted a set of local background genes (LBGs) as a control group in the following analyses (Wang *et al*., 2019). We validated our 103 ARGs using multi-omics data, including functional genomic characteristics and transcriptomics, as well as proteomic profiles generated from diverse AD transgenic mice models.

### Multi-omics validation of network-predicted risk genes in AD

#### A significant disease module formed by ARGs in the human interactome

AD involves a complex, polygenic, and pleiotropic genetic architecture. We found that 103 ARGs formed significantly connected subgraphs (termed disease module) rather than being scattered randomly in the human protein-protein interactome, consistent with previous disease module analyses that we demonstrated in other multiple complex diseases (Cheng and Desai, 2018, Cheng and Lu, 2019). Specifically, 68.0% of ARGs (70/103, *P*=0.015, permutation test) form the largest connected subnetwork (disease module), in comparison to the same number of randomly selected genes with similar connectivity (degree) as the original seed genes in the human interactome (**Supplementary Fig. S1**). This disease module (**Fig. 2A**) includes 128 PPIs (edges or links) connecting 70 unique proteins (nodes). Network analysis revealed 14 proteins with connectivity higher than 5, the top five of which were *ESR1, PSMC5, MAPK1, PAK1* and *NFKB1*. These same five genes have previously been implicated in AD (Granic *et al*., 2009, Conejero-Goldberg *et al*., 2011, Ma *et al*., 2012, Wang *et al*., 2016). For example, *ESR1* interacts with tau protein *in vivo*, and prevents glutamate excitotoxic injury by Aβ via estrogen signaling (Wang *et al*., 2016). Gene expression analysis shows that PSMC5 was significantly overexpressed in patients carrying APOE4 mutations in comparison to APOE4 wild-type group (Conejero-Goldberg *et al*., 2011). In summary, 103 predicted ARGs comprise a strong disease module in the human interactome.

**Figure 2.**
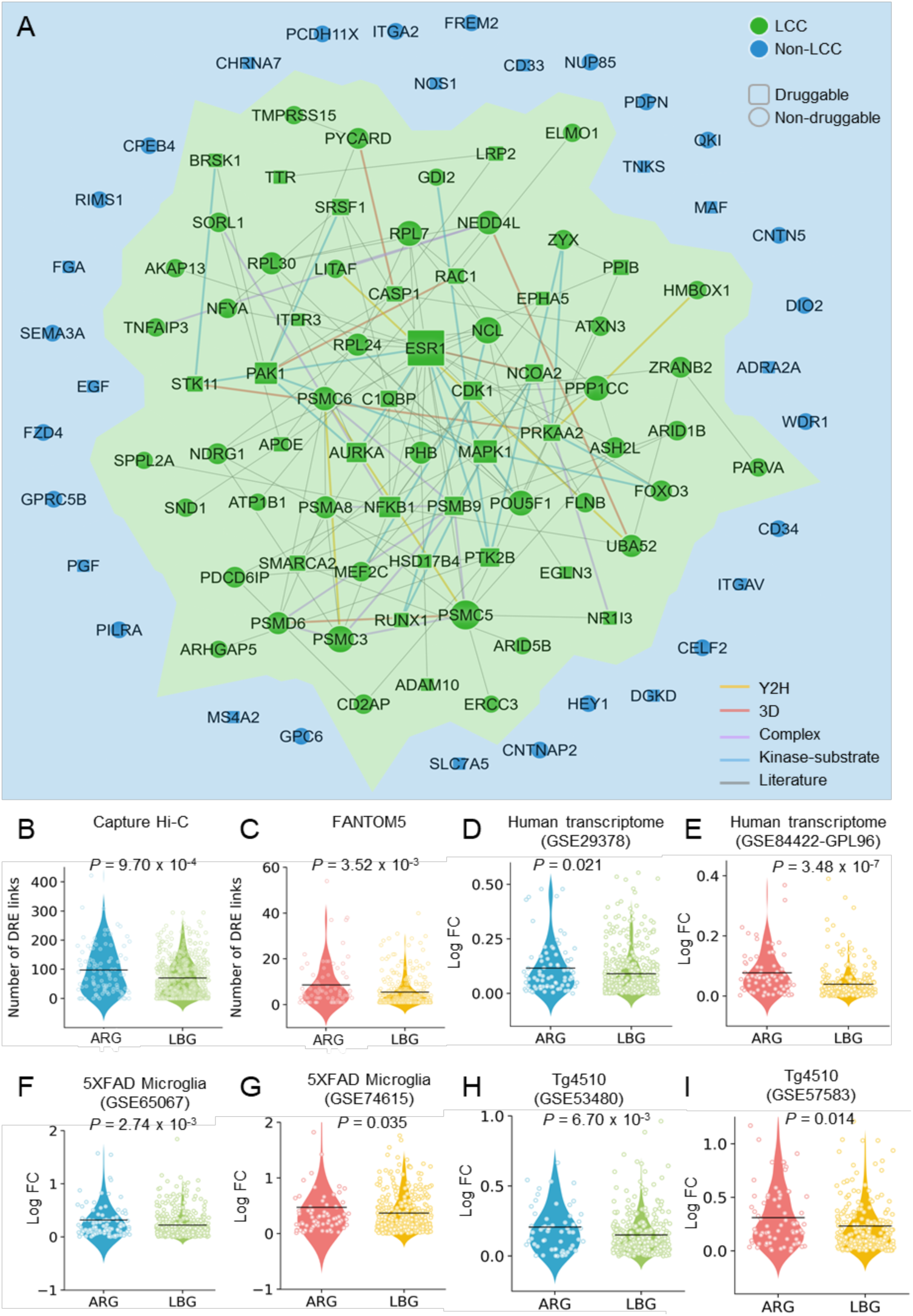
Network-based validation of predicted risk genes for Alzheimer’s disease (AD). **(A)** A subnetwork highlighting disease module formed by predicted AD risk genes (ARGs) in the human protein-protein interactome. This disease module includes 128 protein–protein interactions (PPIs) (edges or links) connecting 70 ARGs (nodes). Larger node size highlighting the high expression level in brain compared to other tissues. **(B-I)** Discovery of genomic features of 103 predicted ARGs implicated in AD. ARGs capture strong distal gene regulatory elements in Hi-C **(B)** and FANTOM5 data **(C)** compared to a set of local background genes (LBGs). (**D-I)** AGRs are more likely differential expression across six transcriptomics datasets: (**D** and **E)** AD patient brain (GSE29378 **[D]** and GPL96 **[E]**), (**F** and **G)** brain microglia cell of 5XFAD mouse model (GSE65067 **[F]** and GSE74615 **[G]**), and (**H** and **I)** brain hippocampus of Tg4510 mouse model (GSE53480 **[H]** and GSE57583 **[I]**). *P* value was computed by one-tail T-test. LCC: Largest Connected Component.

#### ARGs capture more gene regulatory elements

Because a majority of GWAS SNPs lie in non-coding region and exert their function by gene regulation (Zhao *et al*., 2018), we explored the gene regulatory elements of ARGs by testing the hypothesis that the network-predicted risk genes capture more distal regulatory elements (DREs)-promoter connections compared to background. We collected DRE-promoter connection data generated by two technologies: Cap Analysis of Gene Expression (CAGE) from Functional Annotation of the Mammalian Genome 5 (FANTOM5) project and capture Hi-C (see Methods) (Andersson *et al*., 2014, Won *et al*., 2016). Through this, we found that the ARGs are indeed connected to more DREs in both capture Hi-C data (P = 9.70 × 10^−4^, **Fig. 2B**) and FANTOM5 data (P = 3.52 × 10^−3^, **Fig. 2C**).

#### ARGs are more likely to be differentially expressed in AD

We next investigated differential gene expression of ARGs under different pathobiology contexts of AD. Specifically, we measured fold changes of microarray expression levels of ARGs compared to 571 local background genes (LBGs). We found that ARGs were more likely to be differentially expressed in: (i) brain transcriptome of AD patients (**Fig. 2D** and **2E**); and (ii) mouse brain transcriptome from different AD transgenic mouse models (**Fig. 2F-2I)**, including two transcriptomics datasets from 5XFAD mouse brain microglial cells (**Fig. 2F** and **2G**) and two tauopathy-related transcriptomics datasets (Tg4510) from mouse brain hippocampus (**Fig. 2H** and **2I**). We further performed differentially expressed gene enrichment analysis for network-predicted ARGs in AD. We collected bulk RNA-seq data from whole brain tissue or brain microglial cells from two common AD transgenic mouse models (5XFAD and Tg4510) and observed that ARGs were significantly differentially expressed in 5XFAD brain (*P*=0.003), 5XFAD microglial cells (*P*=0.002), and brain microglial of Tg4510 (**Supplementary Table 6**). This suggests that our identified ARGs are potentially involved in the pathobiology of AD.

#### ARGs coding proteins are more likely to be differentially expressed in AD

We further inspected differentially expressed proteins encoded by 103 network-predicted ARGs across 10 published proteomics datasets (see Supplementary Methods). Herein, we evaluated 3 types of AD transgenic mouse models: (a) hAPP model containing APP transgene, (b) 5XFAD model harboring human transgenes for both APP and PSEN1 mutations, and (c) ADLPAPT model carrying three human transgenes (APP, PSEN1 and tau). We found that products of ARGs were significantly differentially expressed in all 3 AD transgenic mouse models (*P* <0.05, Fisher test, **Supplementary Table 6**).

Collectively, we have thus shown that network-predicted ARGs are significantly involved in disease-related functional genomics, transcriptomics, and proteomics, supporting their role as likely causal genes for AD.

### Incorporation of AD multi-omics data to prioritize ARGs

To prioritize ARGs we integrated multi-omics profiles. In total, we incorporated 8 criteria that can be categorized into 5 types of biological evidence: (1) brain-expression specificity (z-score) derived from GTEx database, (2) availability of supportive experimental evidence from the literature and manually curated data from Open Targets database (Koscielny *et al*., 2017), (3) experimentally validated AD genes, (4) differential gene expression, and (5) available drug targets. **Figure 3** shows a global view of 103 ARGs that we validated by these multiple forms of biological evidence in AD. Among 103 ARGs, 89 genes (86.4% [89/103]) satisfy at least one criterion. In addition, 15 ARGs have at least 5 forms of AD-related evidence, including 8 well-known AD genes: *APOE, PTK2B, NOS1, MEF2C, SORL1, EPHA5, ADAM10*, and *CD33*. For the rest of the ARGs, all but BRSK1 had corresponding published literature-derived evidence. For example, PAK1 is a predicted risk gene with 6 criteria of biological evidence: high brain expression specificity (z-score=2.83), supportive experimental evidence from the literature, druggable target data, and, differential expression in human brain of AD patients, microglial cells of 5XFAD mouse model, and brain hippocampus of a tau mouse model (**Fig. 3** and **Supplementary Table 7**). P21-activated kinase 1, encoded by the PAK1 gene, has been implicated in AD (Zhao *et al*., 2006), and recent studies have revealed that inactivation of PAK1 obliterated social recognition without changing amyloid beta (Aβ)/tau pathology, and also exacerbated synaptic impairment and behavioral deficits in mouse models of AD (Arsenault *et al*., 2013, Bories *et al*., 2017).

**Figure 3.**
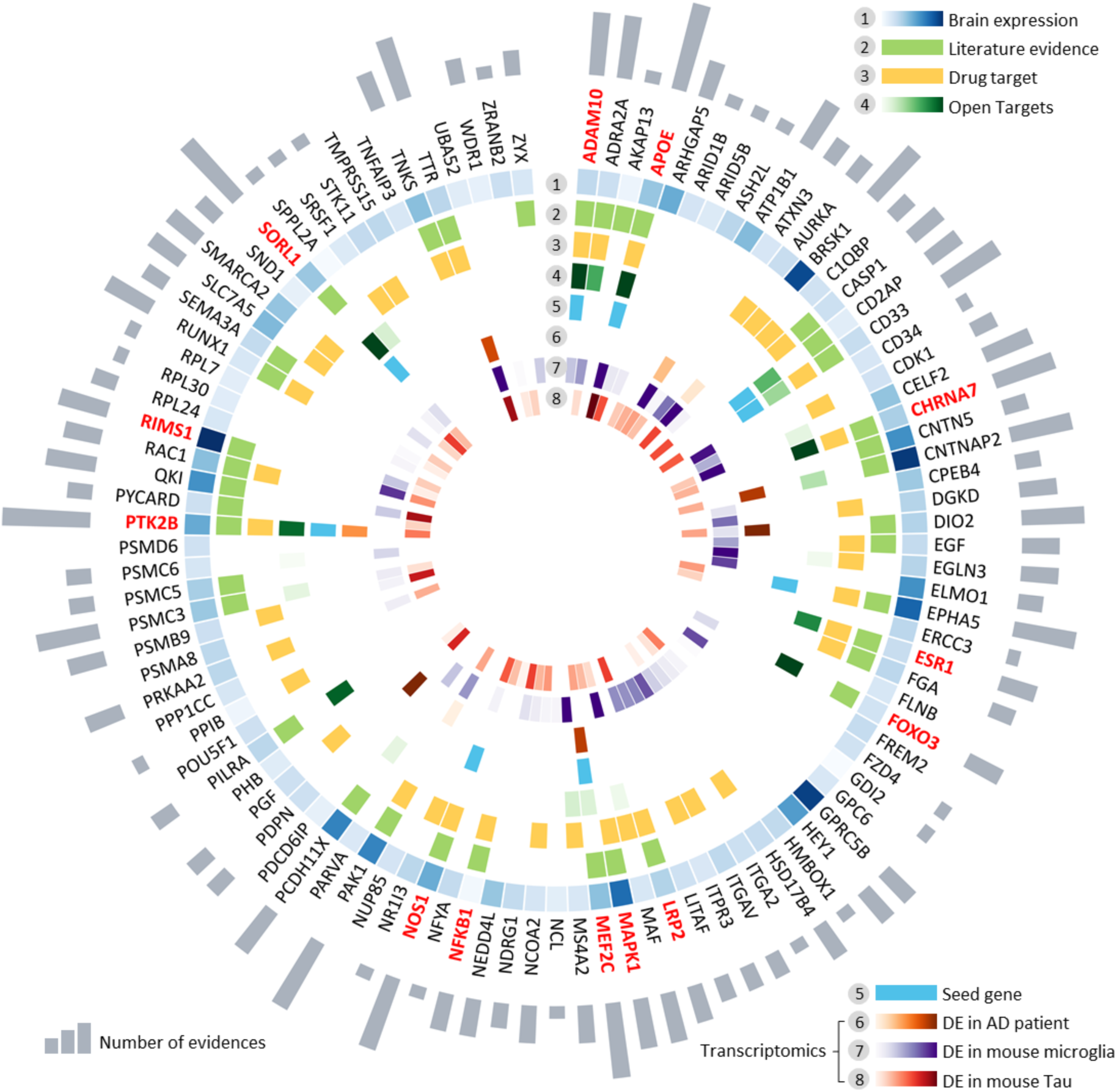
Multi-omics validation of network-predicted risk genes for Alzheimer’s disease (AD). Circle plot shows all 103 predicted AD risk genes validated by multiple-scale biological evidences. In total, 8 types of biological evidences were evaluated: (1) Brain-expression specificity derived from GTEx database (z-score>0 as a high brain-specific expressed gene); (2) literature evidence validation for the gene associated with AD; (3) drug target information; (4) literature-derived experimental data from Open targets database; (5) high quality experimentally validated AD-associated genes; (6) differential expression (DE) in AD patient brains; (7) differential expression in brain microglia cells of 5XFAD mouse model; (8) differential expression in brain hippocampus of Tg4510 mouse model. Gray bar denotes the number of biological evidences. 13 selected risk genes involved in four AD key pathways are highlighted by red: including regulation of neurotransmitter transport, Aβ metabolic process, long-term synaptic potentiation, and oxidative stress.

Among 103 ARGs, we selected 37 (**Supplementary Table 7**) using subject matter expertise based on a combination of factors: (i) high brain-expression specificity, (ii) differential expression in multiple AD transgenic mouse models; (iii) strength of the network-based prediction; and (iv) availability of supportive experimental evidence. To advance disease understanding of network-predicted high-confidence risk genes, we performed biological pathway enrichment analysis using ClueGO plugin in Cytoscape (**Supplementary Table 8**) (Bindea *et al*., 2009). We found 4 statistically significant biological pathways in AD: (a) regulation of neurotransmitter transport, (b) Aβ-related biologic process, (c) long-term synaptic potentiation, and (d) oxidative stress (**Table 1** and **Supplementary Table 9**).

**Table 1.** Network-predicted risk genes involved in four pathobiological pathways of Alzheimer’s disease (AD).

| Gene | Protein | Description |
| --- | --- | --- |
| **Neurotransmitter transport** |  |  |
| MEF2C<sup>a</sup> | Myocyte-specific enhancer factor 2C | MEF2C mRNA expression levels were correlated with AD pathology |
| RIMS1 | Regulating synaptic membrane exocytosis protein 1 | An altered protein expression in RIMS1 during AD pathology |
| **Beta-amyloid-related biologic process** |  |  |
| APOE<sup>a</sup> | Apolipoprotein E | Affect Aβ production, aggregation, and clearance |
| CHRNA7 | Neuronal acetylcholine receptor subunit alpha-7 | Bind to Aβ with very high affinity, providing therapeutic insight into AD |
| SORL1<sup>a</sup> | Sortilin-related receptor | Reduce Aβ generation by trafficking APP away from amyloidogenic cleavage sites |
| ADAM10<sup>a</sup> | Disintegrin and metalloproteinase domain-containing protein 10 | Constitutive α-secretase in the process of amyloid-β protein precursor (AβPP) cleavage |
| LRP2 | Low-density lipoprotein receptor-related protein 2 | rs3755166 polymorphism within LRP2 gene is associated with susceptibility to AD in the Chinese population |
| **Long-term synaptic potentiation** |  |  |
| MAPK1 | Mitogen-activated protein kinase 1 | Beta-amyloid activates the MAPK cascade via hippocampal CHRNA7 |
| PTK2B<sup>a</sup> | Protein-tyrosine kinase 2-beta | An *in vivo* modulator and early marker of Tau pathology. |
| **Oxidative stress** |  |  |
| FOXO3 | Forkhead box protein O3 | Activate BCL2L11 and FASLG to promote neuronal death and aberrant Aβ processing |
| NOS1 | Nitric oxide synthase | Loss of endothelial NOS promotes p25 production and Tau phosphorylation |
| NFKB1 | Nuclear factor NF-kappa-B p105 subunit | Involve in neuroinflammation, synaptic plasticity, learning, and memory implicated in AD |
| ESR1 | Estrogen receptor | Interact with tau protein *in vivo*, and prevent glutamate excitotoxic injury by Aβ through estrogen signaling mechanisms |

<sup>a</sup> denotes that a gene is experimentally validated AD genes. The detailed literature information is provided in **Supplementary Table 9**.

#### Neurotransmitter transport

Specifically, MEF2C and RIMS1, encoding myocyte-specific enhancer factor 2C and regulating synaptic membrane exocytosis protein 1, play key roles in neurotransmitter secretion and synaptic plasticity. MEF2C (rs254776) has been reported in several GWAS studies (Allen *et al*., 2015, Karch and Goate, 2015), and we found significantly lower expression of MEF2C in AD brain (*P* = 1.26 × 10^−3^, one side Wilcoxon test, **Supplementary Fig. 2A**). RIMS1 is a newly predicted ARG, and a recent proteome study from human hippocampus revealed its overexpression in AD (Hondius *et al*., 2016). RIMS1 is significantly overexpressed in 5XFAD mouse microglia (*P* = 4.51 × 10^−3^, one side Wilcoxon test) compared to controls (**Supplementary Fig. 2B**).

#### Beta-amyloid-related biologic process

Five genes (APOE, ADAM10, CHRNA7, SORL1, and LRP2) are associated with beta-amyloid biologic process. Among them, APOE, ADAM10, and SORL1 are well-known AD risk genes, validated by large scale genetic studies and preclinical studies (Fjorback *et al*., 2012, Lambert *et al*., 2013, Kunkle *et al*., 2019). For example, APP cleavage by ADAM10 will produce an APP-derived fragment that is neuroprotective, sAPPα (Peron *et al*., 2018). CHRNA7 (neuronal acetylcholine receptor subunit alpha-7) and LRP2 (low-density lipoprotein receptor-related protein 2) are two newly identified risk genes. There is significantly lower expression of CHRNA7 in the Tg4510 mouse transcriptome (*P* = 4.55 × 10^−4^) compared to controls (**Supplementary Fig. 2E)**. CHRNA7 binds to Aβ with a high affinity (Farhat and Ahmed, 2017). Finally, a previous study showed that the rs3755166 polymorphism within LRP2 is associated with susceptibility to AD in the Chinese population (Wang *et al*., 2011).

#### Long-term synaptic potentiation

Mitogen-activated protein kinase (MAPK1) and PTK2B are two identified risk genes related to long-term synaptic potentiation. Mitogen-activated protein kinase 1, encoded by MAPK1 gene, is highly expressed in brain tissue (z-score = 3.51). The MAPK1 cascade can be activated by Aβ via alpha7 nicotinic acetylcholine receptors (Dineley *et al*., 2001), and significantly lower expression of MAPK1 was found in Tg4510 mice (*P* = 8.27 × 10^−3^) compared to controls (**Supplementary Fig. 2F**). PTK2B, a well-known AD gene with high expression in brain (z-score = 1.62), was identified as an early marker and *in vivo* modulator of tau pathology (Dourlen *et al*., 2017), by mediating Aβ-induced synaptic dysfunction and loss (Salazar *et al*., 2019).

#### Oxidative stress

Oxidative stress is a prominent hypothesis in the pathogenesis of AD (Jiang *et al*., 2016). Here we found four network-predicted ARGs (FOXO3, NOS1, NFKB1, and ESR1) that were associated with regulation of oxidative stress. FOXO3 encoding Forkhead box protein O3 transcription factor, is a direct substrate of CDK5. FOXO3 activates several genes (e.g. BCL2L11 and FASLG) to promote neuronal death and aberrant Aβ processing (Shi *et al*., 2016). Significantly lower expression of FOXO3 was found in 5XFAD mouse microglia (*P* = 8.62 × 10^−4^) compared to controls (**Supplementary Fig. 2H**). NFKB1, encoding transcription factor nuclear factor kappa B (NF-κB), is implicated in oxidative stress, synaptic plasticity, and learning and memory (Snow and Albensi, 2016).

Taken together, these findings suggest that our network-predicted ARGs are involved in diverse pathobiological pathways of AD. However, experimental validations are warranted for several newly predicted ARGs.

### High druggability of network-predicted ARGs

To date, most disease genes generated from GWAS findings are undruggable (Okada *et al*., 2014). For example, a recent study revealed that none of approved and investigational AD drugs target products (proteins) of GWAS-derived genes in AD (Kwok *et al*., 2018). We examined whether network-predicted ARGs were more druggable compared to randomly selected proteins from human protein-coding gene background. Based on drug-target networks from 6 commonly used resources (see **Methods**), we obtained 2,866 potential druggable proteins for FDA-approved or clinically investigational drugs. Surprisingly, we found that 41 out of 103 predicted ARGs (39.8 %) are known druggable proteins, which is four-fold higher than druggable proteins (*P* = 9.25 X 10^−11^, Fisher test) in the genome-wide human protein-coding gene background. High druggability of network-predicted ARGs offers more candidate targets for therapeutic discovery (such as drug repurposing) in AD. For example, ADRA2A, one of the predicted ARGs, encodes adrenoceptor alpha 2A receptor. ADRA2A is a potential target of clozapine (Philibin *et al*., 2009), an atypical antipsychotic drugs. Long-term clozapine treatment reduces Aβ deposition and improves cognitive impairment in an AD transgenic mice model (Choi *et al*., 2017). NR1I3, encoding the nuclear receptor constitutive androstane receptor (CAR), is a potential drug target activated by the lipid-lowering drug simvastatin (Kobayashi *et al*., 2005). Simvastatin was reported to significantly reduce levels of Aβ *in vitro* and *in vivo* (Yamamoto *et al*., 2016, Li *et al*., 2018). In summary, network-predicted ARGs showed higher druggability compared to traditional GWAS-based analysis approaches. We next examined opportunities for drug repurposing by integrating findings from ARGs with the human protein-protein interactome network.

### ARGs offer candidate targets for Alzheimer’s drug repurposing

We have shown that network-predicted ARGs are related to the known pathobiology of AD and offer potential druggable targets, prompting us to examine opportunities for AD therapeutic discovery. We hypothesized that for a drug with multiple targets to be beneficial for treating a disease, its target proteins should be within or in the immediate vicinity of the corresponding disease module (**Fig. 2A**) in the human interactome network. To examine the potential application of ARGs on AD drug repurposing, we applied a network proximity approach (Cheng and Desai, 2018) to quantify the interplay between AD modules from ARGs and drug targets in the human interactome network. We used the cutoff of Z score (Z < -1.5) to select network-predicted repurposable drugs in AD. After exclusion of nutraceutical drugs, metal drugs, and radioactive diagnostic agents, 130 drug candidates were obtained. We then systematically retrieved the published anti-AD clinical, *in vitro/in vivo* reported data for the 130 predicted drugs. In total, 25 had corresponding preclinical or clinical evidence for potential application to AD (**Supplementary Table 10**). **Figure 4** shows the molecular mechanisms of the 25 predicted drug candidates with published experimental or clinical evidence for AD. These drugs are classified into 6 categories according to Anatomical Therapeutic Chemical classification (ATC) codes: musculoskeletal systems [n=6], genito urinary and hormones [n=5], cardiovascular [n=3], alimentary tract and metabolism [n=3], respiratory system [n=2], and others [n=6].

**Figure 4.**
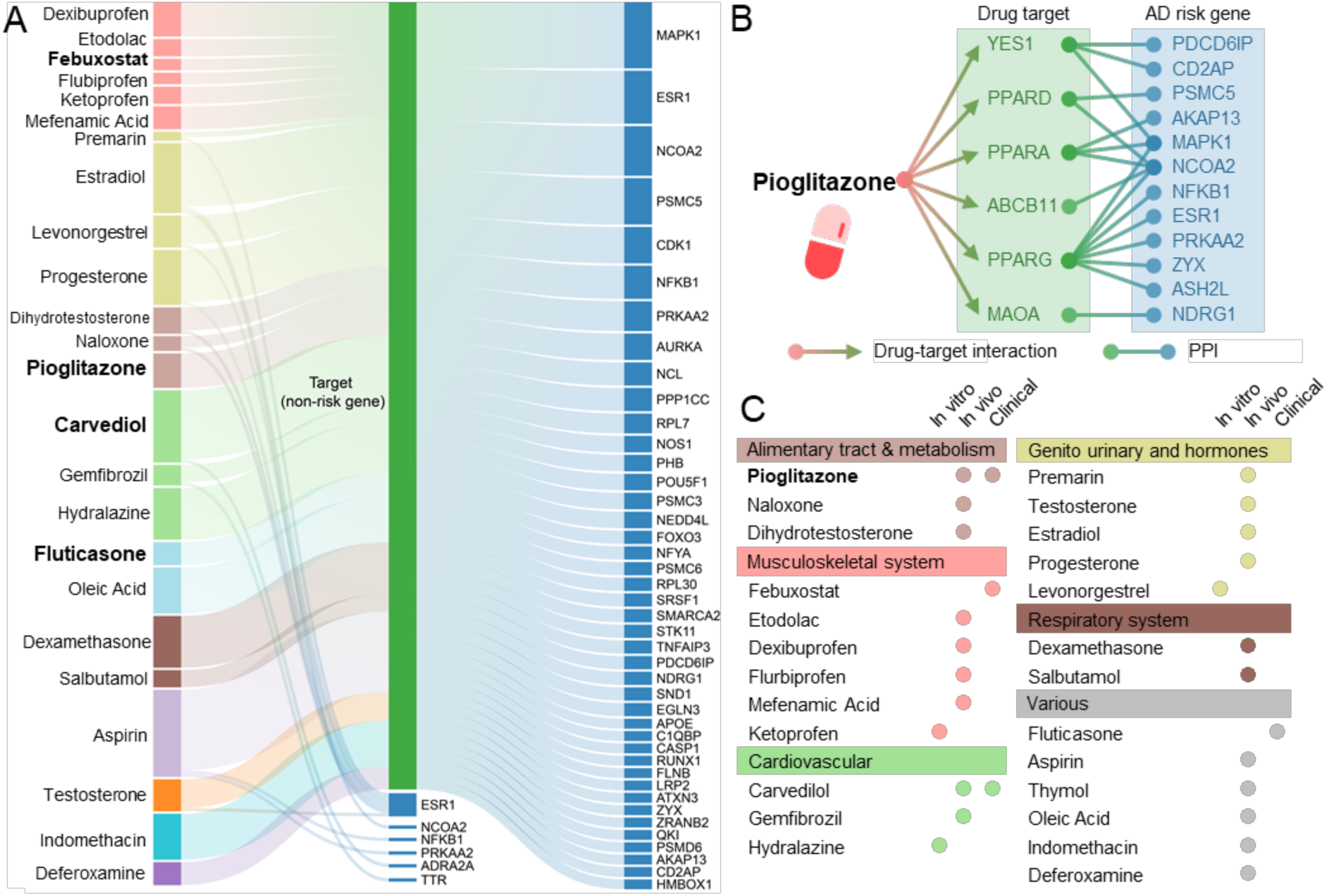
Risk gene-informed drug repurposing for Alzheimer’s disease (AD). (**A**) A Sankey diagram illustrates a global view of 25 repurposable drug candidates with published evidences for AD. These drugs are linked to their physical binding targets or neighborhood proteins derived from network-predicted AD risk genes. (**B**) Network proximity analysis measures the network distance between disease module and drug targets in the human interactome. A subnetwork indicates the molecular mechanism of pioglitazone implicated in AD, which targets six physical binding proteins of which neighborhoods are 12 predicted AD risk genes. (**C**) Drugs are grouped by their first-level Anatomical Therapeutic Chemical Classification (ATC) codes. The drugs with known anti-AD clinical status, *in vitro* and *in vivo* mouse model published data are given. Several approved drugs (pioglitazone, carvedilol, febuxostat and fluticasone) with anti-AD clinical evidence are highlighted.

Among them, we found 4 predicted drugs having known AD clinical evidence (Galimberti and Scarpini, 2017, Lehrer and Rheinstein, 2018), including pioglitazone [NCT02913664], carvedilol [NCT01354444], febuxostat (Singh and Cleveland, 2018), and fluticasone (Lehrer and Rheinstein, 2018). Pioglitazone, an FDA approved drug for T2D, has a significant network proximity (Z = -1.64) with the ARGs. **Figure 4** shows that pioglitazone targets six proteins by connecting with 12 neighborhoods of ARGs.

Several clinical trials conducted with pioglitazone to treat AD. A phase II study (NCT00982202) shows no statistically significant difference between controls and patients with mild to moderate AD (Geldmacher *et al*., 2011). However, another study showed that pioglitazone was associated with cognitive and functional improvement, as well stabilization of AD in 42 diabetic patients (Sato *et al*., 2011). Many of these studies were conducted in populations without biological confirmation of AD by biomarkers and in some case (e.g., the TOMMOW study; NCT01931566), the dose of pioglitazone was substantially lower than that used in clinical practice for the treatment of diabetes. The available clinical trial data do not exclude a beneficial effect of pioglitazone on AD. Except for the TOMMOW study that was conducted in cognitively normal at-risk individuals, other trials have examined symptomatic patients that address a question different from the risk-reduction interrogation we prosecuted. For these reasons, we chose pioglitazone to test the drug user’s relationship with AD outcomes using state-of-the-art pharmacoepidemiologic analysis of large-scale patient data.

### Network-predicted pioglitazone usage reduces risk of AD in patient data

We selected pioglitazone, identified by the network proximity measure (closest), to assess the drug user’s relationship with AD outcomes (**Supplementary Table 11**) by analyzing 7.23 million patients from the Medicare supplemental database (see Methods). Two comparison analyses were conducted to evaluate the predicted association based on individual level longitudinal patient data and state-of-the-art pharmacoepidemiologic methods. These included: (1) pioglitazone (n = 101,650) versus matched control population (n = 410,184), and (2) pioglitazone versus glipizide (a T2D drug, n = 191,656). **Table 2** summarizes the patient data for pharmacoepidemiologic analyses. For each comparison, we estimated the un-stratified Kaplan-Meier curves, conducted by both propensity score stratified (n strata = 10) log-rank test and Cox model. After 6 years of follow-up, pioglitazone significantly reduces risk of AD compared with match control population (*P =* 3.97×10^−4^, hazard ratio (HR) = 0.895, 95% confidence interval [CI] 0.841-0.951, **Fig. 5A** and **5C**). Importantly, propensity score matching cohort studies confirms that pioglitazone is associated with a reduced risk of AD in comparison to glipizide (HR =0.921, 95% CI 0.861-0.983, *P* = 0.0146, **Fig. 5B** and **5C**). Thus, two comparison analyses support our network-based prediction.

**Table 2.** Description of the dataset for the state-of-the-art pharmacoepidemiologic analysis in Alzheimer’s disease.

| Drug | Sample Size | # of AD | Female (%) | Age (Mean) | Geographical location (%) |  |  |  |  | T2D (%) | HTN (%) |
| --- | --- | --- | --- | --- | --- | --- | --- | --- | --- | --- | --- |
|  |  |  |  |  | Northeast | North Central | South | West | NA |  |  |
| Pioglitazone | 101,650 | 1,244 | 38.4 | 73.6 | 17.7 | 28.4 | 35.6 | 17.3 | 0.9 | 51.6 | 34.4 |
| Glipizide | 191,656 | 3,048 | 44.8 | 74.9 | 21.5 | 27.6 | 31.0 | 18.7 | 0.8 | 59.6 | 43.1 |
| Control | 402,184 | 5,498 | 38.4 | 75.46 | 24.6 | 31.6 | 30.3 | 12.4 | 1.0 | 45.0 | 41.4 |

We estimated the un-stratified Kaplan-Meier curves, conducted propensity score stratified (n strata = 10) log-rank test and Cox model. T2D: type 2 diabetes; HTN: hypertension.

**Figure 5.**
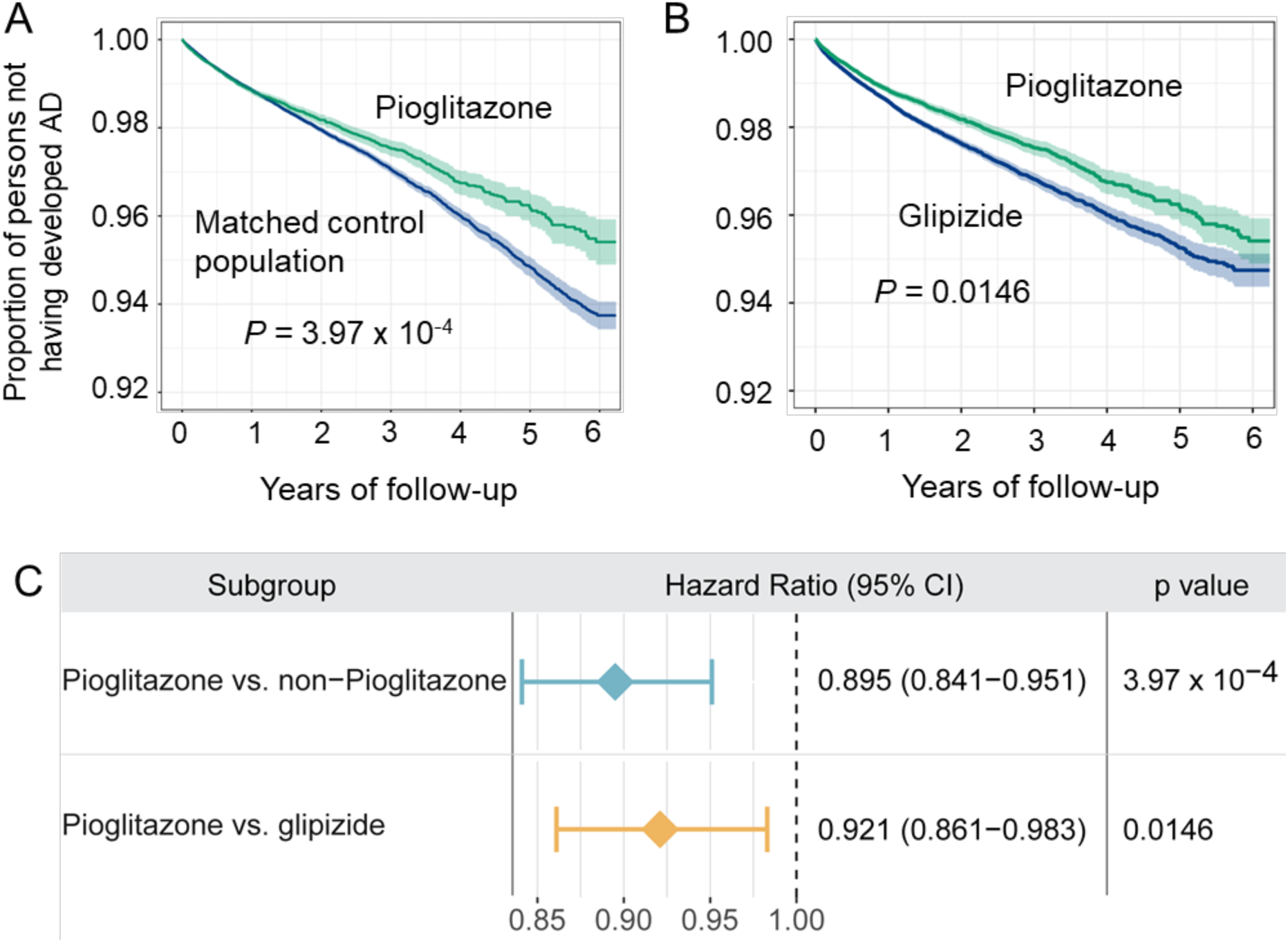
Longitudinal analyses reveal that pioglitazone reduces incidence of Alzheimer’s disease in patient data. Two comparison analyses were conducted including: **(A)** pioglitazone (n = 101,650) vs. matched control population (n = 402,184), and **(B)** pioglitazone vs. glipizide (a diabetes drug, n = 191,656). First, for each comparison, we estimated the propensity score by using the variables described in **Table 2**. Then, we estimated the un-stratified Kaplan-Meier curves, conducted propensity score stratified (n strata = 10) log-rank test and Cox model. **(C)** Hazard ratios and 95% confidence interval (CI) for two cohort studies. Using propensity score stratified survival analyses, non-exposures were matched to the exposures (ratio 4:1) by adjusting the initiation time of pioglitazone, enrollment history, age and gender, and disease comorbidities (hypertension and diabetes).

### *In vitro* observation of pioglitazone’s mechanism-of-action in AD

**Figure 5** reveals that pioglitazone significantly reduces risk of AD in patient-based data. To investigate its mechanism-of-action in AD, we performed a network analysis through integration of drug targets and ARGs into the brain-specific PPI network (see **Methods**). Network analysis shows that pioglitazone potentially targets two tauopathy-related proteins (GSK3β and CDK5) in AD (**Fig. 6A**). RNA sequencing data from the GTEx database (GTEx Consortium, 2015) suggests that GSK3β and CDK5 are highly expressed in brain tissue. Accumulating studies suggested that inhibition of GSK3β and CDK5 activity is a potential therapeutic strategy for AD (Mazanetz and Fischer, 2007).

We next examined pioglitazone’s mechanism-of-action on human microglia HMC3 cells. Firstly, to assess the potential cell cytotoxicity, HMC3 cells were treated with pioglitazone at various concentrations (0.03 µM to 100 µM) for 48 h, and cell viability was determined by MTT method. As presented in **Fig. 6B**, pioglitazone at 0.03 µM to 10 µM did not affect cell viability, revealing low toxicity in human cells. Thus, these optimized concentrations of pioglitazone (≤ 10 µM) were used in subsequent experiments. As shown in **Fig. 6C**, phosphorylation of GSK3β and CDK5 were significantly increased after LPS treatment (1 µg/mL for 30 min) in HMC3 cells. Pre-treating with pioglitazone significantly reduced phosphorylated GSK3β and CDK5 in a dose-dependent manner (**Fig. 6D** and **6E** and **Supplementary Fig. 3**). Altogether, these data suggest that pioglitazone may offer potential benefits for patients with AD by reducing activation of GSK3β and CDK5.

**Figure 6.**
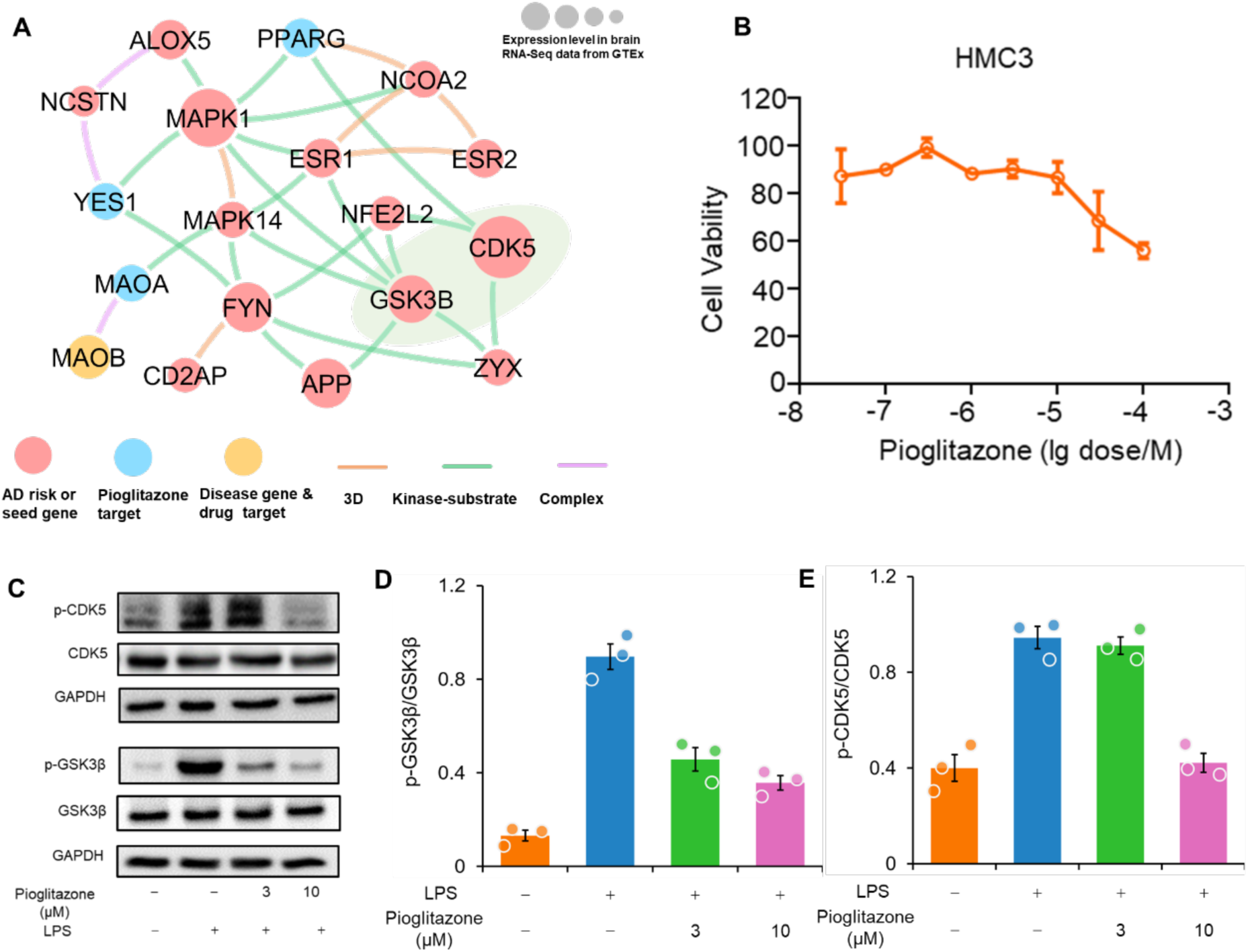
Experimental validation of pioglitazone’s proposed mechanism-of-action in Alzheimer’s disease (AD). **(A)** Network analysis highlighting the inferred mechanism-of-action for pioglitazone in AD. The potential molecular mechanisms of pioglitazone were inferred through integration of known drug targets and predicted AD risk or AD seed genes into brain-specific co-expressed protein-protein interactome network (see the Methods). Node size indicates the protein-coding gene expression level in brain compared with other 31 tissues from GTEx database (GTEx Consortium, 2015). Larger size highlighting the high expression level in brain compared with other tissues. We excluded the literature-derived protein-protein interactions; **(B)** Effects of pioglitazone on the cell viability of HMC3 cells. HMC3 cells were treated with indicated concentrations of pioglitazone for 48 h and cell viability was determined using MTT. Data are represented as mean ± SEM (n = 3) and each experiment was performed at least three times in duplicate. **(C)** Effects of pioglitazone on LPS-induced activation of GSK3β **(D)** and CDK5 **(E)** in human microglia HMC3 cells. HMC3 cells were pre-treated with pioglitazone and followed LPS treatment (1 µg/mL, 30 min). The total cell lysates were collected and subjected to Western blot analysis. Quantification data represent mean ± s.d. of two independent experiments.

## Discussion

AD risk involves a complex polygenic and pleiotropic genetic architecture (Long and Holtzman, 2019). Traditional reductionist paradigms overlook the inherent complexity of human disease and often led to treatments that are inadequate or have important adverse effects (Greene and Loscalzo, 2017). Understanding AD from the point-of-view of how cellular systems and molecular interactome perturbations underlie the disease is the essence of network medicine (Fang *et al*., 2019). In this study, we argue that in the case of AD, cellular networks gradually evolve throughout disease initiation and progression, leading to progressive shifts of local and global network properties and systems states. It is these shifts that underlie the pathogenesis of AD. Genome alterations such as amplification, deletion, and mutation are primary events in AD. However, such events can be selected in human cells only if they encode the appropriate perturbations in the interactome network and systems properties of the affected cells. Therapeutic interventions need to be designed to deal with perturbations of AD systems properties, and have little to do, functionally speaking, with genetic and genomic events alone (Swarup *et al*., 2019). Herein, we have proposed a genome-wide and population-based drug repurposing framework, which integrates genetic findings, functional genomics, drug-target networks, and the human protein-protein interactome, along with large-scale population-based validation and *in vitro* mechanistic observations in human microglia cells.

In total, we identified 103 ARGs by utilizing our recently developed Bayesian model selection method (Wang *et al*., 2019). Human interactome network analysis reveals that ARGs form a statistically significant disease module. Functional genomics enrichment analysis shows that ARGs harbor more gene regulatory elements in the human genome. In addition, large-scale transcriptomics and proteomics data analyses imply that ARGs are more likely to be differentially expressed in human AD brain and multiple AD transgenic mouse models (**Fig. 2**). These comprehensive observations suggest that ARGs potentially capture pathobiological pathways of AD (**Fig. 3**). Importantly, drug-target network analysis shows a 4-fold higher druggability compared to the known drug targets in the human genome.

A previous study showed that few products (proteins) of GWAS-derived closest genes could be applied for therapeutic discovery (Kwok *et al*., 2018). Several factors may account for this. First, the reported significant loci occupy only a small proportion of heritability and provide limited information about underlying AD biology (Deming *et al*., 2017). Second, many genome-wide significant loci lie in noncoding regions, and genes closest to index SNPs may not represent causal genes of AD (Zhang and Lupski, 2015). Thus, systematic identification of likely causal genes from GWAS findings using network approaches is a crucial step for understanding AD pathobiology and offers potential candidate targets for new therapeutic development as presented in this study.

Network-based drug repurposing from ARGs findings predict 4 highly repurposable drugs for AD, including pioglitazone (NCT02913664), carvedilol (NCT01354444), febuxostat and fluticasone. Carvedilol, an FDA approved drug for hypertension that blocks the beta adrenergic receptor, significantly attenuates brain oligomeric β-amyloid level and cognitive deterioration in two independent AD mice models (Wang *et al*., 2011). A propensity-matched analysis has suggested that a daily dose of 40 mg febuxostat can reduce the risk of dementia in older adults (Singh and Cleveland, 2018). Fluticasone is a non-steroidal anti-inflammatory drug (NSAID), and a recent study showed that long term use of fluticasone reduces incidence of developing AD (Lehrer and Rheinstein, 2018).

Pioglitazone, a U.S. FDA-approved anti-T2D drug, was reported to restore energy metabolism and reduce Aβ levels in the brain of APP/PS1 mice (Chang *et al*., 2019). A previous clinical study has shown that pioglitazone improves cognition and regional cerebral blood flow in patients with mild AD accompanied with T2D (Sato *et al*., 2011). However, large-scale population-based validations have not been conducted to confirm the relationship of drug use with AD outcomes in the real-world patient data. In this study, by combining network-based prediction and population-based validation, we found that pioglitazone potentially reduced risk of AD in large-scale patient database (**Fig. 5**). In addition, *In vitro* mechanistic observations (**Fig. 6**) reveal that pioglitazone significantly downregulates expression of CDK5 and GSK3β in human microglia cells, mechanistically supporting network and population-based findings. However, a phase II study (NCT00982202) shows no statistically significant differences between controls and patients with mild to moderate AD for pioglitazone (Geldmacher *et al*., 2011). One possible explanation is that pioglitazone reduces risk of AD only in patients with pre-existing diabetes or that pioglitazone may have its effects before symptoms occur but not in more advanced patients. Thus, our findings suggest that larger clinical trials and additional mechanistic studies may be necessary to clarify pioglitazone’s action in AD prevention in both a broad population and a well-defined sub-population.

Our network methodology presented here has several strengths. First, it contributes to identification of high-confidence likely causal genes, followed by multi-omics data validation, network-based drug repurposing investigation, large-scale patient data analysis, and *in vitro* mechanistic observation in human microglial cells. This work illustrates translation of GWAS findings to pathobiology and therapeutic development in AD. Second, the large patient-level longitudinal data ensures that our analyses integrate real-world patient evidence to test the drug’s efficiency in AD risk reduction.

Potential weaknesses of this work should be acknowledged. First, catalogs of genetic variants from GWAS that influence human disease traits are far from complete, and this deficiency may affect the accuracy of identification of ARGs. Incompleteness of human interactome data and potential literature bias may influence performance of our network methodology. Our choice of validation models (e.g., specific type of transgenic mice models; specific cell types such as microglia) may have influenced our results. Detailed clinical information is missing for health insurance claims data regardless of high-dimensional covariate adjustment. This limits our ability to test the effects of pioglitazone on subpopulation of AD patients such as those with mild AD. In addition, although our dataset contains a geographically diverse population of commercially-insured Americans seniors, the results are not representative of individuals who are not commercially-insured or uninsured. Finally, the phenotyping algorithms (e.g., using the ICD codes) may not capture all AD cases. Thus, this approach may need to be re-applied on a regular iterative basis as datasets are expanded, in order to offer maximum utility.

In summary, this study presents a powerful network-based methodology to translate GWAS findings to emerging therapeutic discovery by exploiting multi-omics, drug-target network, and the human protein-protein interactome, along with large-scale population-based and cell model-based validation. Approaches such as ours may minimize the translational gap between genetic discoveries and therapeutic development in AD and other complex diseases and assist in identifying urgently needed new therapies

## Data Availability

All data used in this study are publicly available as stated in the manuscript.

## Funding

This work was supported by the National Heart, Lung, and Blood Institute of the National Institutes of Health (NIH) under Award Number K99 HL138272 and R00 HL138272 to F.C.

## Author Contributions

F.C. conceived the study. J.F., P.Z., and Q.W. performed experiments and data analysis. Y.Z., R.C., C.W.C, B.Z., B.L., S.J.L., A.A.P., L.L. and J.C., interpreted the data analysis. F.C., J.F., P.Z., and Q.W. drafted the manuscript. F.C., A.A.P., L.L., and J.C. critically revised manuscript. All authors gave final approval of the manuscript.

## Notes

### Competing Interest Statement

The authors have declared no competing interest.

